# Unreliability in Simulations of COVID-19 Cases and Deaths Based on Transmission Models

**DOI:** 10.1101/2024.02.02.24302123

**Authors:** Hideki Kakeya, Makoto Itoh, Yukari Kamijima, Takeshi Nitta, Yoshitaka Umeno

## Abstract

Two papers authored by the same research group were published in academic journals in October 2023, both of which simulate counterfactual COVID-19 cases and deaths using transmission models. One paper estimates that the COVID-19 cases and deaths from Feb 17 to Nov 30, 2021 in Japan would have been as many as 63.3 million and 364 thousand respectively had the vaccination not been implemented, where the 95% confidence interval is claimed to be less than 1% of the estimated value. It also claims that the cases and deaths could have been reduced by 54% and 48% respectively had the vaccination been implemented 14 days earlier. The other paper estimates that the number of cases in early 2022, Tokyo would have been larger than the number of populations in the age group under 49 in the absence of the vaccination program. In this paper, we reexamine the results given by these papers to find that the simulation results do not explain the real-world data in Japan including prefectures with early/late vaccination schedules. The cause of discrepancy is identified as low reliability of model parameters that immensely affect the simulation results of case and death counts. Leaders of public healthcare are required to discern the reliability and credibility of simulation studies and to prepare for variety of possible scenarios when reliable predictions are not available.

## Introduction

In October 2023, two papers on counterfactual simulations of COVID-19 cases and deaths using transmission models were published in two different academic journals. The first paper is titled “Evaluating the COVID-19 vaccination program in Japan, 2021 using the counterfactual reproduction number” [1] and the second paper is titled “Assessing the COVID-19 vaccination program during the Omicron variant (B.1.1.529) epidemic in early 2022, Tokyo” [2].

The first paper claims that the cumulative numbers of COVID-19 infections and deaths in Japan from Feb 17 to Nov 30, 2021 are estimated to be 63.3 million (95% confidence interval [CI] 63.2–63.6) and 364 thousand (95% CI 363–366) respectively in the absence of vaccination, where the actual numbers of reported infections and deaths were 1.2 million and 10 thousand respectively, which means that the vaccination program in Japan reduced mortality by more than 97%. This paper also claims that the cases and deaths could have been reduced by 54% and 48% had the vaccination been implemented 14 days earlier.

The second paper claims that mass vaccination programs directly and indirectly prevented 8.5 million infections (95% CI: 8.4–8.6) during the sixth wave caused by the Omicron variant strains BA.1 and BA.2 from January to May of 2022 in Tokyo, which had a population of 13.8 million. Specifically, this paper estimates that the number of cases in the age group under 49 would have been larger than the number of populations in Tokyo aged under 49 in the absence of the vaccination program, which means that not a small portion of young populations could have been infected with COVID twice during the five-month period.

The simulation results by these papers, especially those by the first paper, were covered and reported by many of the mainstream media in Japan [3-5], partially because of the fame of the corresponding author of these papers, Nishiura, who played a leading role as an advisor to the Japanese government during the COVID-19 pandemic.

While the media coverage of these papers was quoted by some medical doctors to justify the vaccination program against COVID-19 in Japan, many Japanese citizens have cast doubt on these papers, for the numbers of simulated cases and deaths are too enormous to believe.

In this paper, we reexamine the credibility of the parameters Kayano et al. used in the simulations [1,2] by checking the cited data and comparing the simulation results with the actual statistics, including prefectural data where vaccination programs were administered with different time schedules. We also discuss the implication of the result for public health policy making.

### Transmission Models

The two papers on counterfactual simulations of COVID-19 cases and deaths are based on an extended version of the SIR model. In the paper [1], the effective reproduction number, which is interpreted as the average number of secondary cases in age group *a* generated by a single primary case in age group *b* at calendar time *t*, is given by

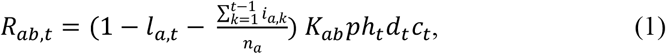

where *l*_*a,t*_ denotes the immune fraction in age group *a* at calendar time *t*, which is based on the estimated efficacy of vaccines, while 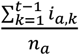 represents the cumulative number of previous infections. Note that left bracketed term in eqn. (1), given by

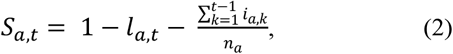

represents the fraction of susceptible populations in age group *a* at time *t*, assuming that none of those previously infected are susceptible. *K*_*ab*_ is a next-generation matrix given by a product of relative susceptibility of age group *a* and the contact matrix between age groups *a* and *b. p* denotes the scaling parameter and *h*_*t*_ expresses the human mobility at time *t. d*_*t*_ represents the increase in transmissibility of the Delta variant compared with earlier variants and *c*_*t*_ expresses the influence of consecutive holidays at time *t* . The confidence interval is calculated by fluctuating the number of infections with a Poisson distribution and applying the maximum likelihood estimation to the scaling parameters related to *p, h*_*t*_, *d*_*t*_, and *c*_*t*_.

In the paper [2], the effective reproduction number is given by

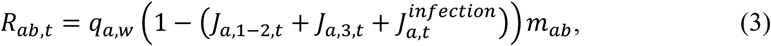

where *q*_*a,w*_ is the weekly scaling parameter in age group *a* in week *w, J*_*a,k,t*_ represents the immune fraction in age group *a* at calendar time *t* attributable to vaccination program *k* (primary series: the first and second doses or booster program: the third dose), 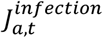 represents the immune fraction owing to immunity that is naturally acquired from infection, and *m*_*ab*_ is a next-generation matrix. The confidence interval is calculated by fluctuating the number of infections with a Poisson distribution and applying the maximum likelihood estimation to *q*_*a,w*_.

### Validation with the Real-World Data

Kayano et al. adjust the parameters to fit the model to all the available data without carrying out validation, which is essential to assess the reliability of the model [6]. It is possible to apply cross-validation to transmission models by dividing the data from each region, part of which is used for training and the rest used for testing. Since the corresponding author of the papers did not respond to our repeated requests to provide us with the source code of their simulator, we use the real-world data in Japan to see whether the results of simulations do not contradict them.

Kayano et al. [1] claim that cases and deaths from Feb 17 to Nov 30, 2021 in Japan could have been reduced by 54% and 48% respectively had the vaccination been implemented 14 days earlier. To confirm the validity of this claim, domestic data including prefectures with early and late vaccination schedules were extracted to see whether early vaccination really contributed to mitigation of damage by COVID.

Prefectures with similar profiles were extracted for fair comparison from [7]. First, the eight most populated prefectures among those ranked in the bottom 12 in the aged (≥ 65) population rate were extracted (Group I). Second, prefectures ranked in the top 17 in the aged population rate were extracted (Group II). Finally, 13 western prefectures (in Chugoku, Shikoku, and Kyushu areas) ranked in the top 25 in the aged population rate were extracted for comparison under similar climate conditions (Group III).

Figure 1 A, B, and C show the rise of the first-dose vaccination rate in the early phase of the vaccination program, while Figure 1 D, E, and F show the relationship between the average vaccination rate *x* (%) between June 1 and August 10 (sampling every two weeks) and the deaths per one million population from COVID *y* between May 12 and November 30. Since the vaccination rate exceeded 1% on May 12 in Japan, the above death count excludes almost all the deaths unrelated to the vaccination program. As vaccination percentage rose by about eight points per 14 days during this time period, COVID death rate should drop by half as the vaccination percentage increases by eight points if the model by Kayano et al. is valid. As shown in the figure, no effect of early vaccination to reduce COVID deaths is observed in all groups.

**Figure 1.**
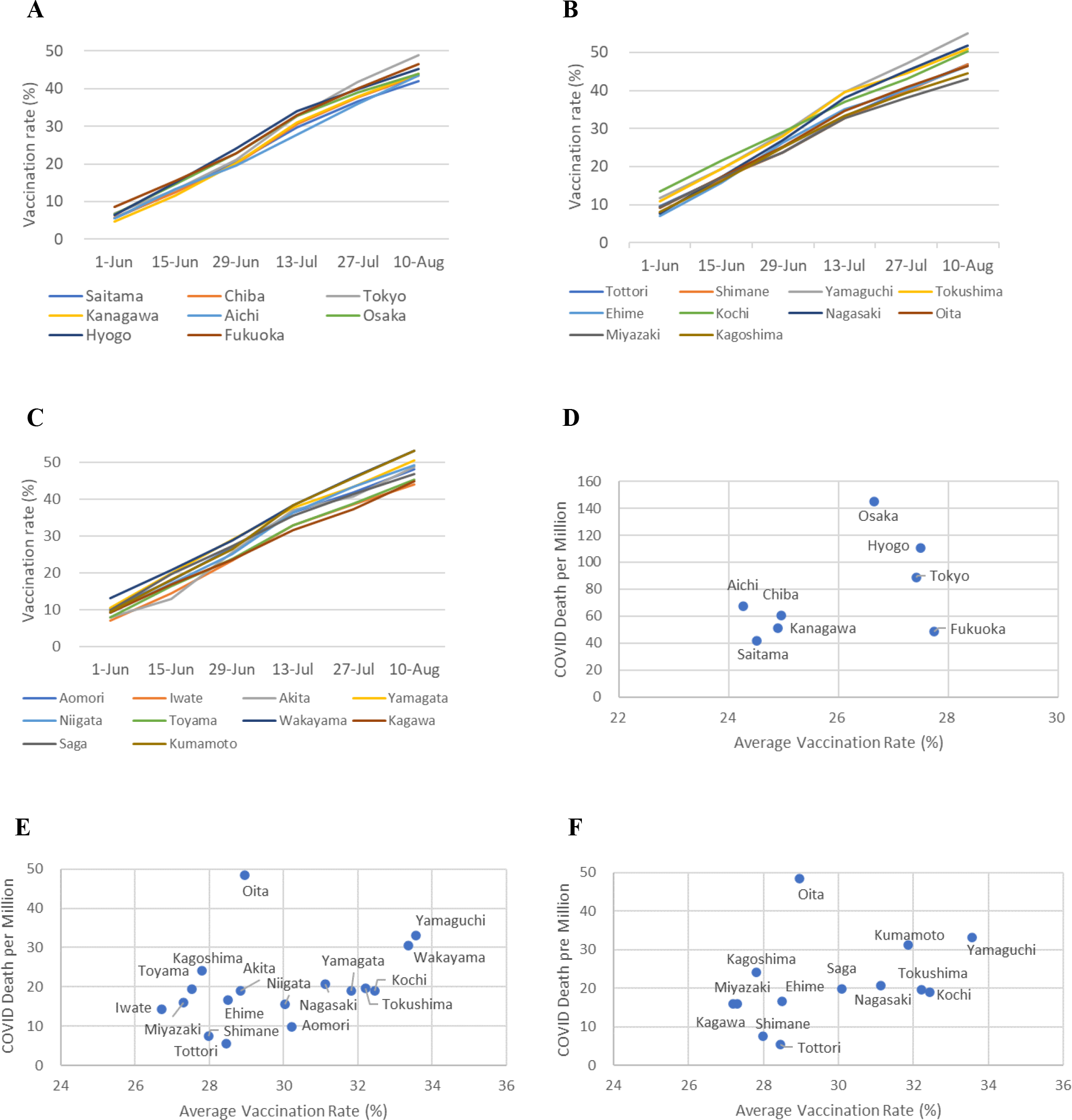
Increase of vaccination rate during the early stage of vaccination program in Group I prefectures (A), in prefectures both in Groups II and III (B), and in prefectures only in one of Groups II or III (C), and relationship between the average vaccination rate from June 1 to August 10 and the deaths from COVID between May 12 and November 30, 2021 in Groups I (D), II (E), and III (F).

To confirm whether the above data can be observed from the model by Kayano et al., we carried out the following test. Kayano et al. [1] claim that COVID deaths would have decreased by 48% if the vaccination program had started 14 days earlier and would have increased by 50% if it had started 14 days later. This prediction includes deaths from February 17 to November 30.

Let the average COVID deaths per million population by February 17, May 11, and November 20 be *D*_1,_, *D*_2,_, *D*_3_ respectively. To reproduce the results by Kayano et al., *y* = *y*_*e*_ = 0.52(*D*_3_ − *D*_1_) − (*D*_2_ − *D*_1_) should hold under 14-day early vaccination and *y* = *y*_*l*_ = 1.5(*D*_3_ − *D*_1_) − (*D*_2_ − *D*_1_) should hold under 14-day late vaccination. Let us assume that the vaccination rate *x* increases by *r* percentage points per 14 days on average. Let us also assume that the curve to predict the death per million population under different vaccination speed passes through the centroid of data (*x, y*) = (*u, v*). Then the curve is supposed to pass through (*u* − *r, y*_*l*_), (*u, v*), and (*u* + *r, y*_*e*_). The prediction model can be obtained by finding a quadratic equation *y* = *ax*^2^ + *bx* + *c* that passes through the above three points.

Based on the parameters calculated above, the differences between the values predicted by the model and the real-world data (the former minus the latter) are plotted for Groups I, II, and II, as shown in Figure 2. As this figure shows, the errors are biased with negative correlation. The result of regression analysis indicates that the correlation is statistically significant in Groups II and III (*p* = 0.0073 and *p* = 0.023), which corroborates the bias in the prediction model by Kayano et al.

**Figure 2.**
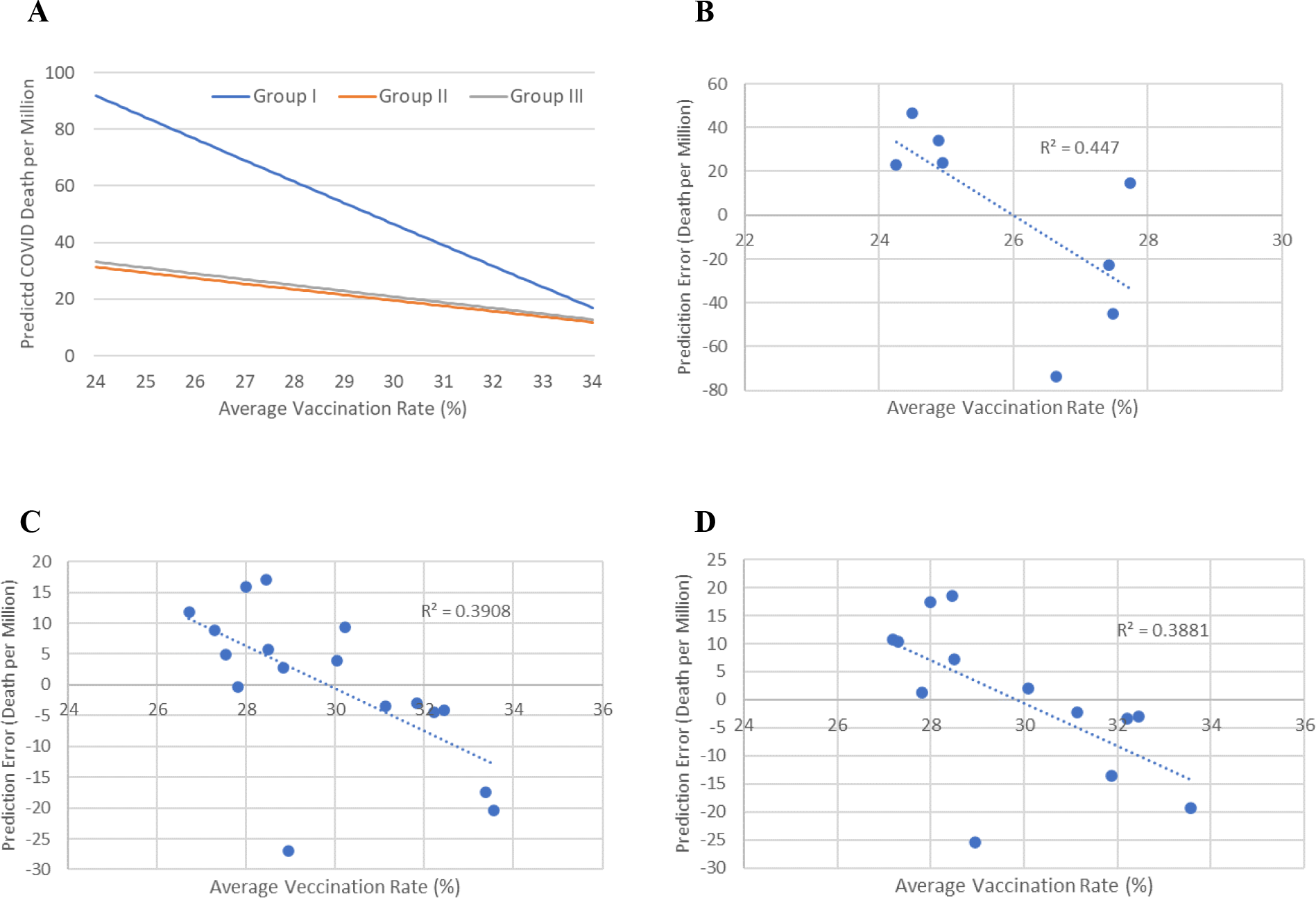
Prediction curve based on the results by Kayano et al. [1] (A) and the differences between the values of the prediction curve and the real-world data in Groups I (B), II (C), and III (D).

### Reliability of Parameters in the Transmission Model

The real-world data strongly indicate that the simulations by Kayano et al. are not reliable. To identify what was wrong in their model, the detail of their transmission model was scrutinized.

Kayano et al. changed *d*_*t*_ in eqn. (1) to reflect the increase of infectivity in the Delta variant. During the surge of the Delta variant, the sensitive fraction given by *S*_*a,t*_ in eqn. (2) decreased as the vaccinate rate increased, which is reflected by the increase of *l*_*a,t*_ in eqn. (2). Therefore, *d*_*t*_ has to be large enough to exceed the effect of vaccination to generate a large peak. Kayano et al. presumes that the Delta variant is 1.5 times infectious than the Alpha variant.

The problem is that the infection pattern can be reproduced with various combination of parameter values. The same infection curve is generated when *R*_*ab,t*_ does not change. Therefore, when *d*_*t*_ is smaller, the same infection curve is reproduced by adjusting *l*_*a,t*_ so that the value *S*_*a,t*_ *d*_*t*_ may remain constant. Estimated large number of cases and deaths are generated in the simulation of late or no vaccination scenarios by eliminating or delaying the rise of the term *l*_*a,t*_ under a large *d*_*t*_, while the estimated number becomes small when *l*_*a,t*_ and *d*_*t*_ are both small. Therefore, the reliability of the parameters *l*_*a,t*_ and *d*_*t*_ is crucial to see that of the counterfactual simulations.

If one of the parameters *l*_*a,t*_ and *d*_*t*_ is reliable and fixed, the other is fixed automatically to fit the real data. Therefore, the model can be reliable if one of them is based upon solid evidence. However, the evidence Kayano et al. rely on for the estimation of *l*_*a,t*_ and *d*_*t*_ is extremely weak. As for the estimation of *l*_*a,t*_, Kayano et al. refer only to a single previous study [8], which does not provide enough information on the waning of vaccine protection. COVID-19 Forecasting Team carried out a systematic review and meta-analysis on the waning of vaccination protection against infection, symptomatic disease, and sever disease, where the range of confidence interval is extremely wide [9].

On the other hand, Kayano et al. refer to three previous studies for estimation of maximum *d*_*t*_ [10-12], while only one of them can be the basis of their assumption that the Delta variant is 1.5 times more infectious than the Alpha variant [11], where the Alpha variant is estimated to be 1.29 times more infectious (CI: 1.24-1.33) and the Delta variant is estimated to be 1.97 times more infectious (CI: 1.76-2.17) than the original Wuhan strain. Since the confidence interval is both around ±10%, the effect of the estimation error of *d*_*t*_ should be taken seriously, for it works exponentially in the SIR model, which means 10% error makes large difference after a long time span. Kayano et al. claim that their estimated value has a confidence level of less than 1% without considering the errors in estimation of *l*_*a,t*_ and *d*_*t*_, which is fatal as an academic study to be published in a scientific journal. It is also noteworthy that the above parameters used for simulations of transmission in Japan all derive from the data outside Japan with little regard to ethnic factor, which does not meet the standard of epidemiological study either.

The second paper by Kayano et al. [2] claims that the number of COVID cases averted by booster in Tokyo from January to May of 2022 is larger than the population in each age group [13] under 49, as shown in Table 1. Notably, the numbers of averted cases in 20-29 age group is about 1.4 times the population of the age group, meaning that more than 40% of the youth in their 20s could have been infected with COVID twice during the five-month time period, which is quite implausible. By looking into the assumption by Kayano et al. carefully, this simulation result comes from their assumption that the protection against infection naturally acquired from infection 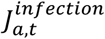 is as weak as and as fast to wane as that given by vaccination, which does not reflect the real-world data [9].

**Table 1.** Estimation by Kayano and Nishiura [2] on the number of COVID cases averted by booster in Tokyo from January to May of 2022 and the actual population in Tokyo in each age group.

| Age group (years) | Cases averted by booster (95% CI) | Population |
| --- | --- | --- |
| 0-9 | 1,425,061 (1,418,927-1,431,610) | 1,050,377 |
| 10-19 | 1,268,196 (1,261,495-1,274,982) | 1,060,726 |
| 20-29 | 2,411,696 (2,398,716-2,424,445) | 1,714,279 |
| 30-39 | 2,392,823 (2,378,472-2,407,955) | 1,905,423 |
| 40-49 | 2,340,664 (2,323,896-2,357,358) | 2,179,987 |
| 50-59 | 1,649,600 (1,634,595-1,664,511) | 2,022,404 |
| 60-69 | 894,544 (883,907-904,551) | 1,375,781 |
| 70-79 | 634,301 (626,165-642,737) | 1,422,047 |
| $\geq 80$ | 596,883 (589,711-603,884) | 1,063,908 |

## Discussion

A simulation without cross-validation or any other validation with the real-world data does not meet the standard of academic research. Neither does the confidence interval without taking into account the possible errors in parameters that immensely affect the simulation result. Simulations with unrealistic assumptions should not be used to estimate what would have happened under a counterfactual scenario. We regret that the serious flaws in the papers have been apparently overlooked through the reviewing process.

It is often pointed out that publications of medical journals tend to put too much emphasis on the implications of the paper for the medical community rather than scientific rationality and integrity [14], leading to spread of false or subjective interpretation of the facts. Indeed, regardless of the aforementioned unreliability, the papers by Kayano et al. have been used to justify the vaccination program in Japan after the publications, accompanied by a large-scale coverage by the mainstream media, which was cited repeatedly by medical doctors in Japan who promote further vaccination.

Japan is known for its outstanding number of COVID vaccination per capita, while as many as 5965 injuries including 453 death-related after vaccination have been accepted by the government to apply relief system for injury to health with vaccination by Jan 26 of 2024 [15]. A recent study shows statistical bias in the sex of those who died within 10 days after vaccination between the age groups under 65 and 65 or over, indicating the influence of vaccination on the occurrence of death [16]. In 2022, excess deaths amounted to 113,000 in Japan [17], some of which are suspected to have been caused by the vaccination as in other countries, which will be evaluated in future studies. Since the results of the simulation here could be used for justification of those sacrifices by unduly inflating the number of people saved by the vaccination, the undependability of the simulation results should deserve attention not only of scientific communities but also of the general public.

Nishiura, the corresponding author, has been working as an advisory committee member of the government during the COVID pandemic. On July 29 of 2021, just before the Tokyo Olympics, he said in a media interview [18] that the surge of the Delta variant might not subside even if human mobility was decreased as low as that under the first state of emergency declaration. A month later, he said in an interview with the same media [19] that he had predicted the case of infections would exceed ten thousand in the latter half of August, which was obviously wrong.

On May 4 of 2023, just before the update of COVID-19 categorization from category 2 (the same with tuberculosis, avian flu, et al.) to category 5 (the same with seasonal flu), Nishiura expressed a deep concern over the revision of COVID-19 categorization in a media interview and said unprecedented number of elderly people all over Japan would get infected and die eventually [20] if the countermeasure was loosened against COVID, which actually did not happen.

Thus, predictions on the surge of infection by Nishiura in the media have failed repeatedly, which has been witnessed by the citizens in Japan. As discussed in this paper, the number of cases and deaths simulated by an SIR model and its extension is largely influenced by a small error in the effective reproduction number, which is almost impossible to estimate with a high accuracy. Therefore, quantitative predictions or estimations of infection cases and deaths are virtually impossible. Insisting what is unpredictable predictable is a clear violation of scientific integrity. Publications of unreliable predictions by scientists lead to unpreparedness for unpredicted situations. Leaders of public healthcare should ignore unreliable scientific predictions and prepare for variety of possible scenarios when reliable predictions are unattainable.

It is also imperative that scientists take responsibilities for their predictions when they intend to use them for political purposes [21], like objections to Tokyo Olympics or to category revision of infectious disease. Integrity of medical science in general is under jeopardy [22.23], which should be rectified to have individual and group accountability [24].

## Data Availability

All the data used for the study are available from the corresponding author upon request.

## Data availability statement

All the data used for the study are available from the corresponding author upon request.

## Author contributions

HK carried out the whole data analysis. YK supervised this study from an epidemiological point of view. TN supervised this study from an immunological point of view. MI and YU supervised this study from a mathematical point of view.

## Acknowledgement

The authors thank Profs. Hiroshi Tauchi, Yoshihisa Matsumoto, and Takashi Nakamura for comments and suggestions on the manuscript.

